# Economic effects of a country-level tobacco endgame strategy: a modelling study

**DOI:** 10.1101/2023.03.16.23287269

**Authors:** Driss Ait Ouakrim, Tim Wilson, Samantha Howe, Philip Clarke, Coral Gartner, Nick Wilson, Tony Blakely

## Abstract

**Background:** Aotearoa-New Zealand (A/NZ) is the first country to pass a comprehensive commercial tobacco endgame strategy into law. Key components include the denicotinisation of smoked tobacco products and a major reduction in tobacco retail outlets. Understanding the potential long-term economic impacts of these measures is important for government planning.

**Methods:** A tobacco policy simulation model that evaluated the health impacts of the A/NZ Smokefree Action Plan was extended to evaluate the economic effect of the new measures from both Government and citizen perspectives. Estimates were discounted at 3% per annum and presented in 2021 purchasing power parities US$.

**Findings:** The modelled endgame policy package generates considerable growth in income for the A/NZ population with a total cumulative gain by 2050 amounting to US$31 billion. From a government perspective, the policy results in foregone tobacco excise tax revenue with a negative net financial position estimated at US$11.5 billion by 2050. In a sensitivity analysis considering future changes to labour workforce, the government’s cumulative net position remained negative by 2050, but only by US$1.9 billion.

**Interpretation:** Our modelling suggests the Smokefree Aotearoa 2025 Action Plan is likely to produce substantial economic benefits for the A/NZ population, and modest impacts on government revenue and expenditure related to the reduction in tobacco tax and increases in aged pensions due to increased life expectancy. Such costs can be anticipated and planned for and might be largely offset by future increases in labour force and the proportion of 65+ year olds working in the formal economy.

**Funding:** This study was funded by a grant from the Australian National Health and Medical Research Council (GNT1198301)

**Research in Context:** *Evidence before this study:* Multiple countries have set targets to achieve a commercial tobacco endgame. Most simulation modelling studies have evaluated ‘traditional’ tobacco control interventions (e.g., tobacco excise tax increases, indoor smoking bans, smoking cessation health services). Very few have modelled the economic effects of endgame strategies. We searched PubMed with no language restrictions for articles published from 1 January 2000 to 8 February 2023 using the following search terms: (smoking[TW] OR tobacco[TW]) AND (endgame[TW] OR eliminat*[TW] OR “phasing out”[TW] OR “phase out”[TW] OR aboli*[TW] OR prohibit*[TW] OR ban[TW] OR “smoke free”[TW] OR “smoke-free”[TW]) AND (model*[TW] OR simulat*[TW]) AND (cost[TW] OR economic[TW]). We identified six economic evaluations of commercial tobacco endgame strategies, including different interventions and cost perspectives. Five studies modelled interventions in the Aotearoa/New Zealand (A/NZ) context and one in the UK. Four studies were conducted from a healthcare system perspective, estimating the costs to the health system associated with tobacco-related diseases. One of these studies additionally estimated ‘non-health social costs’, as the productivity loss resulting from smoking-associated morbidity and mortality. Another study estimated the cost to consumers resulting from a policy in which retail outlets selling tobacco were significantly reduced, considering both the actual cost of a pack of cigarettes and the cost of increased travel to retailers, and the last estimated excise tax revenue to the government resulting from increases to tobacco taxation (compared to no increases to current tobacco tax levels). Of the identified literature, none evaluated the effect of endgame strategies on citizen income nor the fiscal impacts to government revenue and expenditure.

*Added value of this study:* This study evaluates the economic impacts of a recently introduced commercial tobacco endgame legislation in A/NZ. We modelled the economic impacts by 2050 of a policy package that includes the four key measures in the new legislation (i.e., denicotinisation of smoked tobacco products, enhanced antismoking mass media campaigns, 90% reduction in the number of tobacco retail outlets, and a smoke-free generation law that bans sale of tobacco to anyone born after 2008). The analysis presents both a government and citizen perspective. The government fiscal impacts extend beyond health system expenditure to also include differences between business as usual (BAU) – i.e., no endgame strategy – and endgame scenarios in excise tax revenue, goods and services tax (GST) revenue, income tax revenue, and superannuation expenditure. A net government position is also calculated. The citizen perspective estimates the impact of the policy on population income and savings that may result from reduced tobacco consumption. Our model projects large economic gains for consumers from the tobacco endgame package resulting from a sharp reduction in smoking prevalence, morbidity and mortality. For the A/NZ Government, the policy is projected to result in reduced healthcare costs, and increased income tax and GST revenue. These gains are offset by increased superannuation payments resulting from a greater number of individuals living past the age at which superannuation is provided to all citizens (65 years in A/NZ and described in this article as “retirement age” for simplicity), as well as large reductions in excise tax revenue.

*Implication of all the available evidence:* Our findings support previous evidence indicating that ambitious tobacco control policies can produce large heath and economic benefits. Our model suggests that a commercial tobacco endgame strategy is likely to result in a large revenue transfer to the benefit of the A/NZ population. An endgame approach moves beyond the BAU model of incremental policy change to a deliberate strategy to permanently reduce tobacco smoking to minimal levels within a short timeframe. A logical result of such a strategy is a significant decrease in excise tax revenue for governments. Under the endgame scenario, the net position of the A/NZ Government is likely to be negative due mainly to the foregone excise tax revenue. In a sensitivity analysis of the endgame scenario that takes into account recent projections from Stats NZ of a future larger and older labour force in A/NZ, our model suggests that the net government position might become positive as early as 2036 – less than 15 years after the introduction of the endgame policy.

## INTRODUCTION

Smoking is a leading cause of avoidable morbidity and mortality.^1^ Globally, the annual economic loss due to smoking has been estimated at US$1,436 billion, equivalent in magnitude to 1.8% of the world’s annual gross domestic product (GDP).^2^ In the United States (US) alone, the annual loss in income and unpaid household production due to tobacco consumption has been estimated at $436 billion per annum - equivalent to 2.1 % of the 2020 GDP for that country.^3^

In this context of massive health and economic losses due to tobacco, commercial tobacco endgame strategies are being increasingly proposed and regarded as a viable approach to tackle the tobacco epidemic.^4^ An endgame approach moves beyond the business-as-usual (BAU) model of incremental policy change to a deliberative strategy to permanently reduce tobacco smoking to minimal levels within a short timeframe, or a complete phase out of the commercial tobacco market. The endgame concept is often interpreted as a smoking prevalence of ≤5% in the adult population.^5^ As of early 2023, ten countries (including Aotearoa-New Zealand [A/NZ], England, Scotland, Republic of Ireland, US, Canada, Australia, Sweden, Finland, and Bangladesh) have announced goals to reach the ≤5% target between 2025 and 2040.^6^

Among these countries, A/NZ is the first to pass into law a package of policies aiming to reduce smoking prevalence to ≤5% prevalence before 2030 and to reduce the inequity in smoking rates between the Māori (Indigenous) and non-Māori populations. When operationalised, the Smokefree Environments and Regulated Products (Smoked Tobacco) Amendment Act, which was passed by the Parliament in December 2022,^7^ will reduce the nicotine content of all smoked tobacco products to non-addictive levels, reduce the number of tobacco retail outlets by at least 90%, and ban tobacco sales to anyone born after 2008.^8^ These new policies are likely to be accompanied by enhanced smoking cessation support, grants to engage community groups in activities to achieve the smokefree goal and other health promotion activities. In a recent study we evaluated the potential health impacts of these policies and found that their implementation would deliver large health and equity gains compared to a BAU approach.10. According to our modelling, a combined tobacco endgame policy package would lead to a gain of 594,000 health-adjusted life years (HALYs; 95% uncertainty interval [UI]: 443,000 to 738,000; 3% discount rate) over the remaining lifetime of the 5.08 million A/NZ population alive in 2020.^9^

Despite the unprecedented potential for a commercial tobacco endgame to increase population health and equity, and to reduce healthcare expenditure and lost productivity due to premature death and disability, phasing out commercial tobacco sales often raises concerns about economic impacts on governments from loss of tobacco taxes. In this study, we aimed to quantify the potential economic effects of the Smokefree Aotearoa 2025 Action Plan from both Government and citizen perspectives.

## METHODS

We used a previously published simulation model^9^ developed to evaluate the health impacts of the A/NZ Smokefree Action Plan. Details of the model’s methodology, design, assumptions and epidemiological parameters have been reported elsewhere.^9-12^ Briefly, the simulation is based on the combination of two models: 1) a Markov process simulating the population’s smoking and vaping life history based on seven states (see supplementary Figure S1). Movements between the different states are determined by transition probabilities, which reflect BAU and additional super-imposed effects of the intervention (see below); 2) a proportional multistate lifetable (PMSLT) composed of a main cohort lifetable, which simulates the evolution of A/NZ population from 2020 using projected all-cause mortality and morbidity rates. For this analysis, we evolved the model from a closed-cohort to an open-cohort simulation by including births and migration using projections from Stats NZ (the A/NZ official data agency). In parallel, in the BAU scenario, proportions of the cohort also reside in 16 subsidiary tobacco-related disease lifetables according to prevalence at baseline, and in future years based on BAU disease-specific incidence, case fatality and remission rates (where appropriate e.g., for treated cancers). The tobacco-related diseases in the model are coronary heart disease, stroke, chronic obstructive pulmonary disease (COPD), lower respiratory tract infection (LRTI), and the following cancers: lung, oesophageal, stomach, liver, head and neck, pancreas, cervical, bladder, kidney, endometrial, melanoma, and thyroid.

### Economic outcomes

Table 1 lists the economic input parameters included in the model and their sources. We identified baseline estimates of total population income, total government income tax revenue, goods and services tax (GST) revenue, tobacco excise tax revenue, superannuation expenditure and health expenditure for the year 2021 from the Financial statement of the Government of A/NZ.^13^ Within each disease lifetable, these parameters were allocated by five-year age groups to proportions of the cohort as follows: population income was attached to cohorts aged 20 to 64 years, superannuation payments were attached to cohorts aged 65 years and older, tobacco excise was attached to the proportion smoking. Health expenditure by disease was attached to all cohorts. The model was calibrated to produce values that match the baseline economic parameter estimates after one cycle run (i.e., 2021). Table 2 presents the economic outcomes produced by the model and their calculation method.

**Table 2:** Aggregate differences in economic outputs between tobacco endgame and BAU

| Output | Calculation |
| --- | --- |
| $\Delta$ Population income | Population income<sub>Intervention</sub> – Population income<sub>BAU</sub> |
| $\Delta$ After tax population income | $(\Delta \text{ Population income}) \times (1 - \text{income tax rate}^\dagger)$ |
| $\Delta$ GST revenue | $(\Delta \text{ Population income}) \times \text{GST rate}^\ddagger$ |
| $\Delta$ Income tax revenue | $\text{Income tax rate}^\dagger \times \Delta \text{ population income}$ |
| $\Delta$ Tobacco excise revenue | $(\text{Number of people who smoke}_{\text{Intervention}} - \text{Number of people who smoke}_{\text{BAU}}) \times \text{Tobacco excise tax rate}^\ddagger$ |
| $\Delta$ Health system expenditure | Total health expenditure<sub>Intervention</sub> – Total health expenditure<sub>BAU</sub> |
| $\Delta$ GST revenue from tobacco sales | $0.15/1.15 \times (\text{Number of people who smoke}_{\text{Intervention}} - \text{Number of people who smoke}_{\text{BAU}}) \times \text{Tobacco expenditure rate}^\ddagger$ |
| $\Delta$ Population expenditure on tobacco | $(\text{Number of people who smoke}_{\text{Intervention}} - \text{Number of people who smoke}_{\text{BAU}}) \times \text{Tobacco expenditure rate}^\ddagger$ |
| $\Delta$ Superannuation expenditure | $(\text{Difference in population aged } 65+^\# \text{ between endgame and BAU}) \times \text{superannuation expenditure rate}^\ddagger$ |
| $\Delta$ Government net position | $\Delta \text{ Income tax revenue} + \Delta \text{ Tobacco excise revenue} + \Delta \text{ GST revenue} - \Delta \text{ Health system expenditure} - \Delta \text{ Superannuation expenditure}$ |

\* Tobacco industry is defined as all producers and retailers – consistent with the calculation method.
<sup>#</sup> Age of superannuation entitlement varies under sensitivity analysis scenario with dynamic retirement age.
<sup>‡</sup> Parameter from Table 1

**Table 1:**
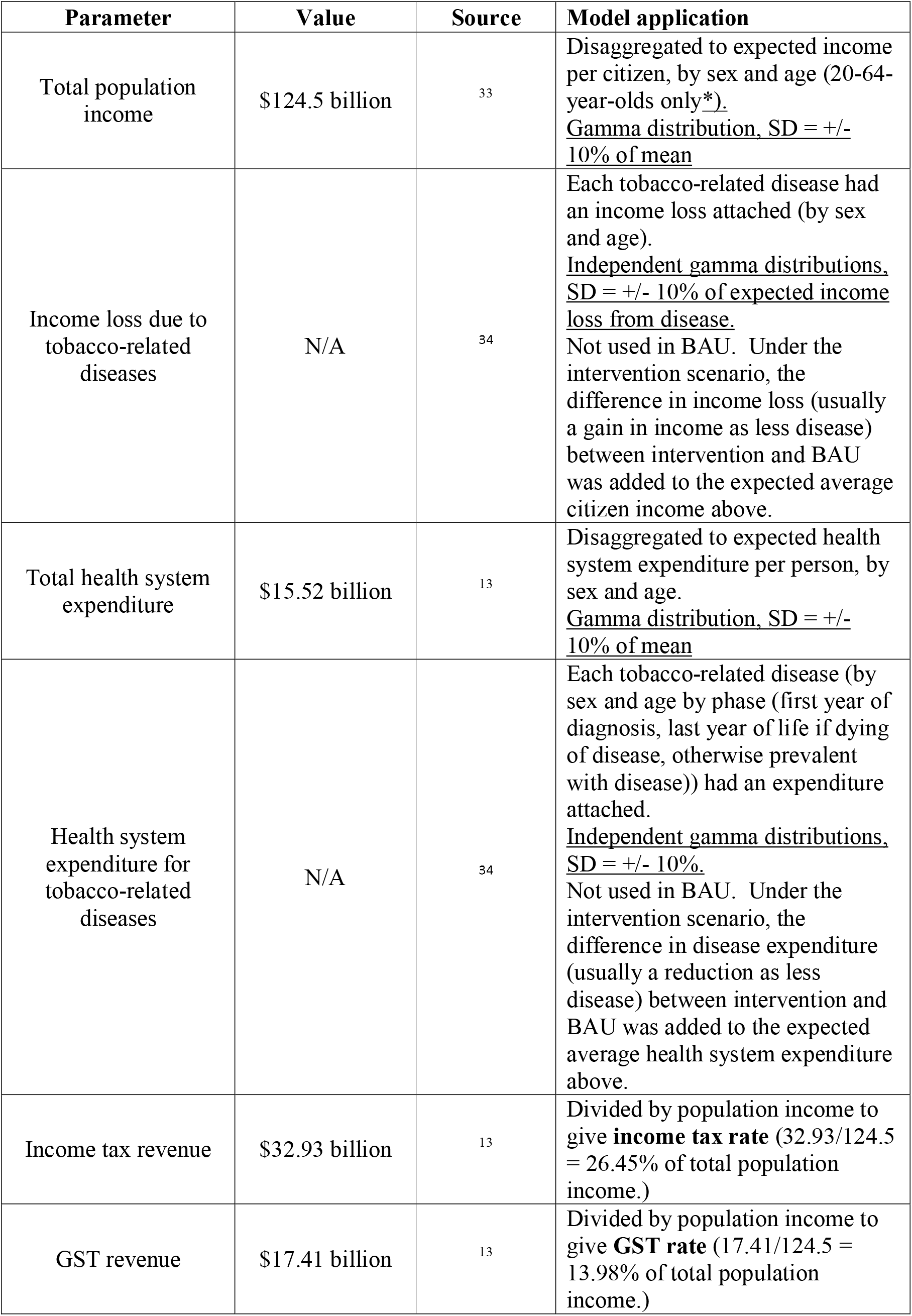
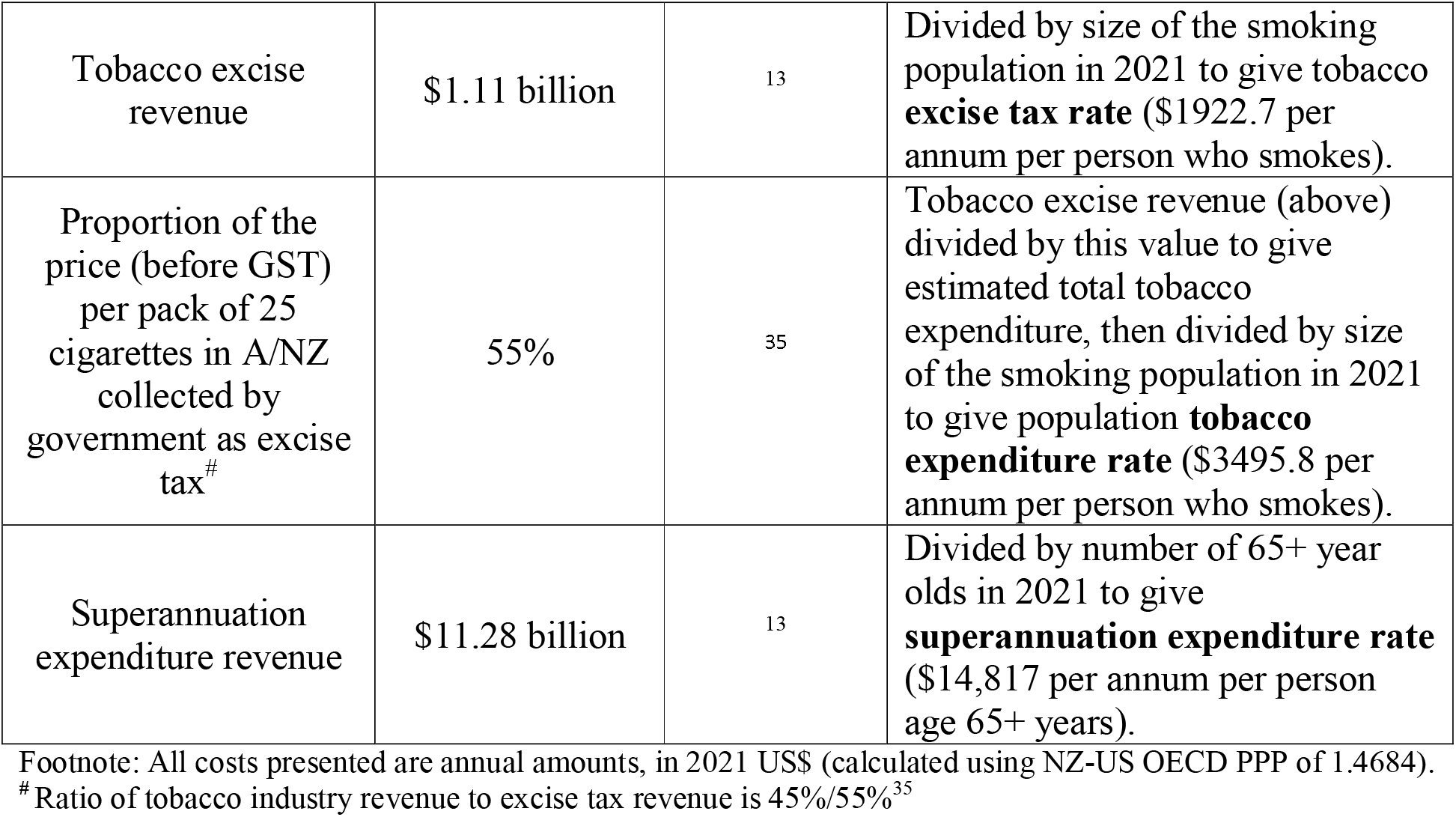
Base-year (2021) annual cost inputs to the modelling, and application within model

| Parameter | Value | Source | Model application |
| --- | --- | --- | --- |
| Total population income | \$124.5 billion | 33 | Disaggregated to expected income per citizen, by sex and age (20-64-year-olds only*).<br><u>Gamma distribution, SD = +/- 10% of mean</u> |
| Income loss due to tobacco-related diseases | N/A | 34 | Each tobacco-related disease had an income loss attached (by sex and age).<br><u>Independent gamma distributions, SD = +/- 10% of expected income loss from disease.</u><br>Not used in BAU. Under the intervention scenario, the difference in income loss (usually a gain in income as less disease) between intervention and BAU was added to the expected average citizen income above. |
| Total health system expenditure | \$15.52 billion | 13 | Disaggregated to expected health system expenditure per person, by sex and age.<br><u>Gamma distribution, SD = +/- 10% of mean</u> |
| Health system expenditure for tobacco-related diseases | N/A | 34 | Each tobacco-related disease (by sex and age by phase (first year of diagnosis, last year of life if dying of disease, otherwise prevalent with disease)) had an expenditure attached.<br><u>Independent gamma distributions, SD = +/- 10%.</u><br>Not used in BAU. Under the intervention scenario, the difference in disease expenditure (usually a reduction as less disease) between intervention and BAU was added to the expected average health system expenditure above. |
| Income tax revenue | \$32.93 billion | 13 | Divided by population income to give **income tax rate** ($32.93/124.5 = 26.45\%$ of total population income.) |
| GST revenue | \$17.41 billion | 13 | Divided by population income to give **GST rate** ($17.41/124.5 = 13.98\%$ of total population income.) |
| Tobacco excise revenue | \$1.11 billion | 13 | Divided by size of the smoking population in 2021 to give tobacco **excise tax rate** (\$1922.7 per annum per person who smokes). |
| Proportion of the price (before GST) per pack of 25 cigarettes in A/NZ collected by government as excise tax<sup>#</sup> | 55% | 35 | Tobacco excise revenue (above) divided by this value to give estimated total tobacco expenditure, then divided by size of the smoking population in 2021 to give population **tobacco expenditure rate** (\$3495.8 per annum per person who smokes). |
| Superannuation expenditure revenue | \$11.28 billion | 13 | Divided by number of 65+ year olds in 2021 to give **superannuation expenditure rate** (\$14,817 per annum per person age 65+ years). |

Footnote: All costs presented are annual amounts, in 2021 US\$ (calculated using NZ-US OECD PPP of 1.4684).
<sup>#</sup> Ratio of tobacco industry revenue to excise tax revenue is 45%/55%<sup>35</sup>

For each simulated year, a population impact fraction (PIF) is calculated for each tobacco-related disease. The generic formula^14^ is:

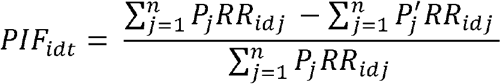

where:

i subscripts each sex by age by ethnic group d subscripts each disease

t subscripts each time step or yearly cycle

j subscripts each category of smoking or vaping (the seven states in supplementary Figure 1, plus 20 additional tunnel states for each of those quitting smoking and/or vaping and people who switched completely from smoking to vaping)

RR is the incidence rate ratio for disease d and smoking-vaping state j, and possible varying by demographics (e.g., by sex and age, but not by ethnic group). (Note the RR does not vary by time step t.)

These PIFs are the percentage change (compared to BAU) in incidence rates for each smoking-related disease, by socio-demographics and year, that are transferred to the PMSLT.

Within each disease lifetable, the endgame intervention is run in parallel to BAU with different disease incidence rates given changes in smoking and vaping prevalence over time (see supplementary Figure S1). Each disease lifetable estimates the difference between intervention and BAU in disease mortality, morbidity, and the modelled economic outputs (Table 2). These differences are calculated at the end of each one-year cycle then added to matching entities in the all-cause or main lifetable.

### Intervention

Intervention effects were reflected in the model through changes in population movements (i.e., transition probabilities) between smoking and vaping states. The endgame policy package considered in the model combines the effects of four separate interventions included in the Smokefree Aotearoa 2025 Action Plan: 1) denicotinisation, 2) enhanced mass media campaign, 3) 95% reduction in the number of tobacco retail outlets; and 4) smoke-free generation. Parameterisations of the individual policies and the combined smokefree policy package are described in Supplementary Table S1. This paper focuses on the combined effect of these interventions if implemented as a single policy package in 2023.

### Sensitivity analysis: dynamic retirement age scenario

The economic outcomes based on transfer payments between Government and citizens included in our model are heavily dependent on the evolution of the labour force in A/NZ. Therefore, our projection of the net government position (i.e., once all the transfers have been considered) is likely to be sensitive to the size and participation of the working-age population. The latest report from Stats NZ National Labour force projections estimates that by 2043 the median size of the labour force in A/NZ will rise by 17.2% compared to 2020. Over the same period, the proportion of the labour force aged 65 years and older is projected to increase from 6% in 2020 to 7-11% in 2043 and 7-15% in 2073.^15^

To test the sensitivity of our model to these labour force evolutions, we developed an alternative endgame scenario with a ‘dynamic’ age of retirement and access to superannuation (i.e., pension payments). Under this scenario, the threshold age increases each year so that the citizen morbidity rate (people who do and do not smoke combined) under the intervention matches the morbidity rate of a 65-year-old under BAU (i.e., without the tobacco endgame intervention). That is, the dynamic scenario is one way to try and capture the contribution that a prevention program such as the A/NZ tobacco endgame legislation might make to the healthiness and thence productivity of the population.

This firstly involved measuring prevalent Years Lived with Disability (pYLDs; measure of average morbidity for a given population) for age 65–70-year-olds, for each year up to 2050, as follows:

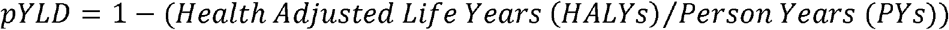

Secondly, for each year, the updated age of superannuation entitlements (i.e., the dynamic retirement age) was calculated as follows:

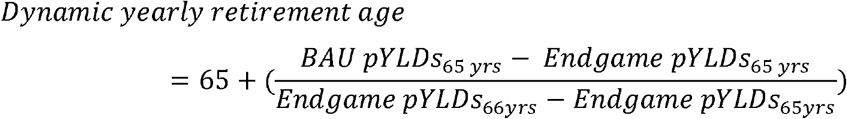

HALYs and Person-Years (PYs) for the above equation were calculated within the PMSLT for both BAU and the alternative endgame scenario.^16^

All scenarios were run 2000 times in Monte Carlo simulation. A 3% discount rate per annum was applied to all economic measures. Undiscounted results are provided in the online supplementary material. Estimates were calculated in 2021 NZ$ then converted to US$ using a 2021 NZ-US OECD purchasing power parity adjustment of 1.4684.

## RESULTS

Figure 1 shows the annual differences in costs between the endgame scenario and BAU. Table 3 presents the cumulative expenditure and revenue estimates.

**Figure 1:**
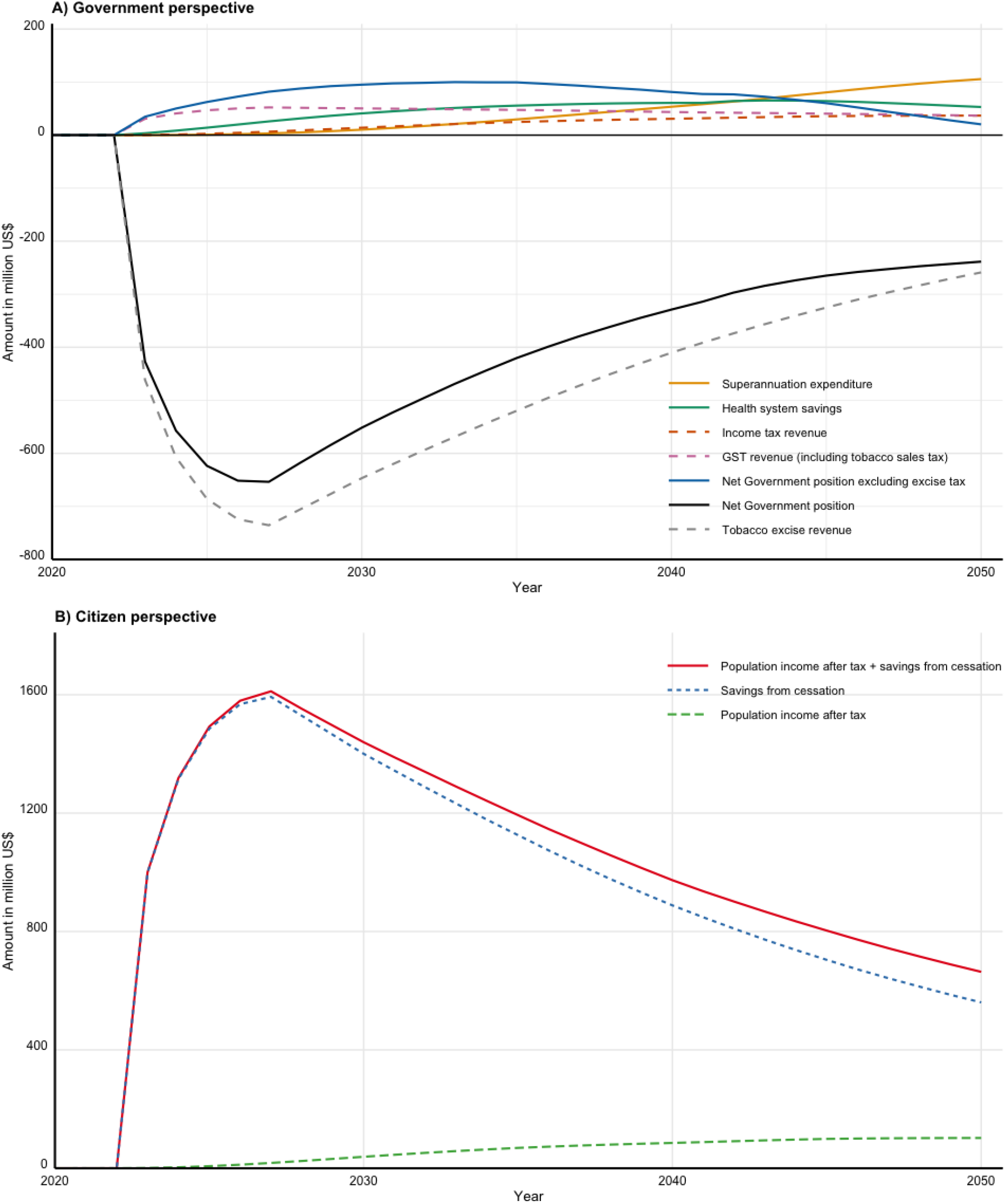
Estimated annual differences in revenue and expenditure (2021 US$; 3% annual discount rate) between the tobacco endgame scenario and BAU.

**Table 3:** Projected changes in cumulative expenditure and revenue due to the Aotearoa-New Zealand’s tobacco endgame strategy compared to BAU (2021 PPP US$ billions; 3% annual discount rate)

| Revenue/expenditure items | by 2030 |  | by 2040† |  | by 2050† |  |
| --- | --- | --- | --- | --- | --- | --- |
|  | Estimate | 95% UI | Estimate | 95% | Estimate | 95% |
| **Government perspective** |  |  |  |  |  |  |
| ***Expenditure*** |  |  |  |  |  |  |
| Health system | -0.18 | (-0.22 to -0.14) | -0.73 | (-0.90 to -0.57) | -1.34 | (-1.70 to -1.02) |
| Superannuation expenditure | 0.03 | (0.02 to 0.04) | 0.35 | (0.28 to 0.43) | 1.18 | (0.93 to 1.44) |
| ***Revenue*** |  |  |  |  |  |  |
| Income tax revenue | 0.05 | (0.04 to 0.06) | 0.30 | (0.24 to 0.35) | 0.65 | (0.52 to 0.77) |
| GST revenue (including tobacco sales tax) | 0.37 | (0.3 to 0.44) | 0.84 | (0.68 to 0.99) | 1.24 | (0.99 to 1.48) |
| Tobacco excise revenue | -5.24 | (-6.16 to -4.24) | -10.35 | (-12.26 to -8.24) | -13.56 | (-16.39 to -10.53) |
| **Net Government position**<br>($\sum \text{revenue} - \sum \text{expenditure}$) | -4.67 | (-5.49 to -3.77) | -8.83 | (-10.52 to -6.96) | -11.51 | (-14.03 to -8.77) |
| **Citizen perspective** |  |  |  |  |  |  |
| Population income after tax | 0.14 | (0.11 to 0.17) | 0.82 | (0.66 to 0.98) | 1.80 | (1.46 to 2.15) |
| Savings from cessation (reduced tobacco expenditure) | 11.35 | (13.35 to 9.19) | 22.41 | (26.55 to 17.83) | 29.36 | (35.49 to 22.80) |
| Population income after tax + savings from cessation | 11.49 | (9.30 to 13.50) | 23.20 | (18.50 to 27.45) | 31.16 | (24.35 to 37.47) |

Note: 2020 NZ\$ converted to 2021 US\$ using NZ-US OECD purchasing power parity of 1.4684.
† i.e., includes estimate to left, as cumulative over time.

For the endgame scenario compared to BAU, health system expenditure savings discounted at 3% per annum are projected to peak in 2044 at US$65m (95% UI: 49 to 83) before decreasing to US$53m (95% UI: 35 to 72) in 2050. The health system is projected to save a cumulative total of US$1.34 billion (b) (95% UI: 1.02 to 1.7) by 2050. Conversely, government expenditure in superannuation benefits will increase by a cumulative total of US$1.18b (95% UI: 0.93 to 1.44), over the same period, due to people living longer.

Population income (after tax) increases on average by US$5m every year after the introduction of the policy (i.e., 2023), reaching US$138m annually (95% UI: 113 to166) in 2050. This represents a projected cumulative income gain of US$1.8b (95% UI: 1.4 to 2.1) by 2050. This increase in income leads to a parallel increase in government income tax revenue.

If money not spent on cigarettes is diverted to other expenditure in the economy, then the effective increase in cumulative disposable income is projected to be US$31.16b (95% UI: 24.3 to 37.4) by 2050. Assuming the increase in disposable income is fully spent in the economy, government GST revenue increases by a cumulative total of US$1.24b (0.99 to 1.48) by 2050.

Annual Government revenue from tobacco excise for the endgame scenario compared to BAU falls rapidly to a maximum of US$735m (608 to 837) less revenue in 2027. The cumulative excise tax revenue foregone by 2050 is US$13.5b (95% UI: 10.5 to 16.4).

The net of revenue and expenditure differences between the endgame and BAU from the Government perspective is dominated by the tobacco excise tax loss. There is a net loss for the Government in every year out to 2050, and a cumulative negative net position of US$11.51b (95% UI: 8.7 to 14.0) by 2050.

Figure 2 and Table 4 show the same results, but for the scenario analysis that sees the age of retirement and eligibility for superannuation increase over time, so the new threshold age has the same morbidity as 65-year-olds in BAU. Under this scenario, the age threshold for entitlement to superannuation becomes 65.2 years in 2030, 65.5 years in 2040 and 65.8 years in 2050. The Government’s net annual position compared to BAU becomes positive by 2037 (Figure 2) – due to changes in income tax revenue and superannuation payments. This scenario still resulted in a net cumulative loss to the Government of US$1.89b (-4.74 to 1.01) by 2050 but is only 14% of the similar loss with a static age.

**Figure 2:**
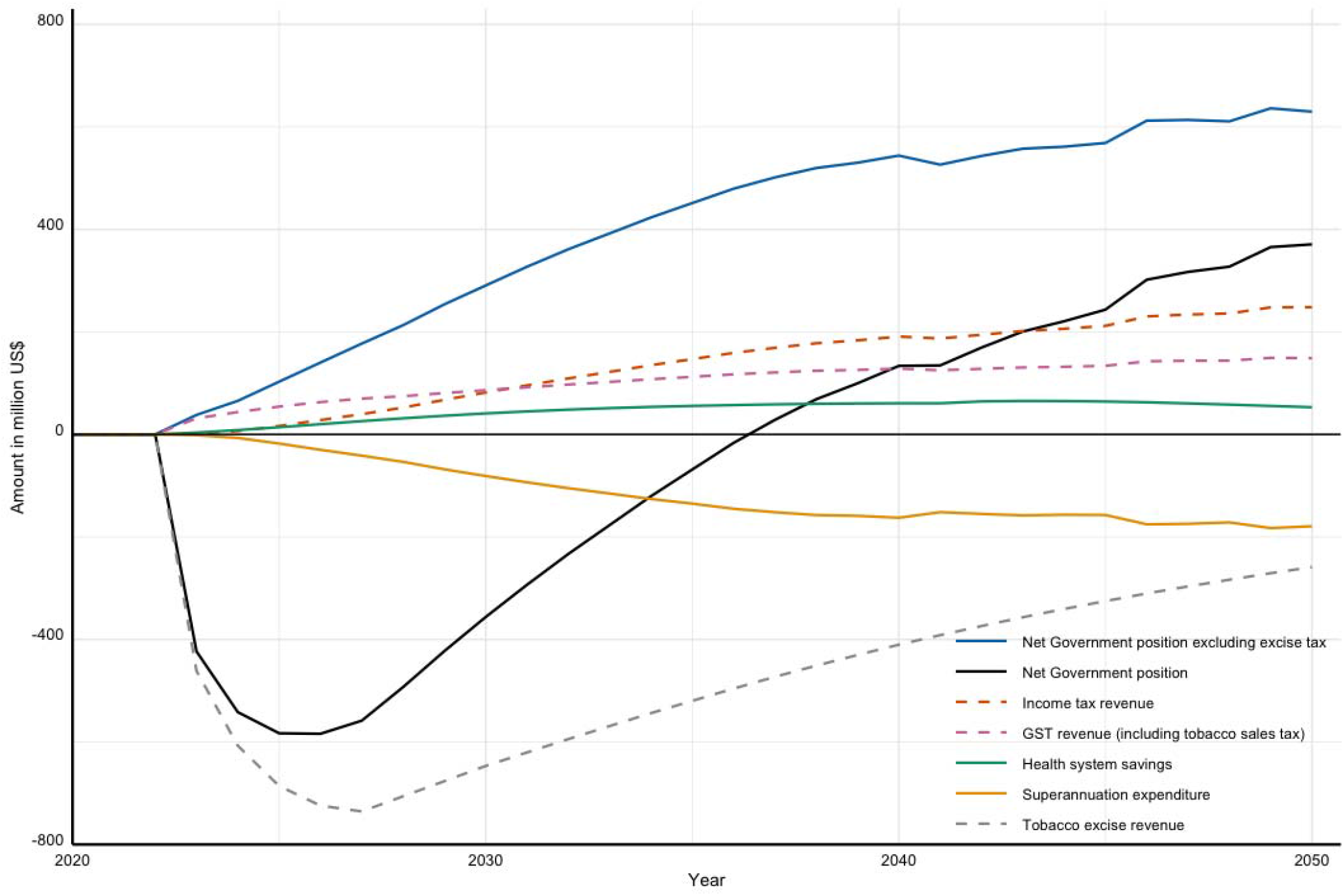
Estimated annual differences in revenue and expenditure (2021 PPP US$; 3% annual discount rate) between tobacco endgame scenario with dynamic retirement age and BAU (Government perspective)

**Table 4:** Projected changes in cumulative expenditure and revenue due to the Aotearoa-New Zealand tobacco endgame strategy compared to BAU using dynamic retirement age (2021 PPP USD billions; 3% annual discount rate)

| Revenue/expenditure items | by 2030 |  | by 2040† |  | by 2050† |  |
| --- | --- | --- | --- | --- | --- | --- |
|  | Estimate | 95% UI | Estimate | 95% | Estimate | 95% |
| **Government perspective** |  |  |  |  |  |  |
| ***Expenditure*** |  |  |  |  |  |  |
| Health system | -0.18 | (-0.22 to -0.14) | -0.73 | (-0.90 to -0.57) | -1.34 | (-1.70 to -1.02) |
| Superannuation expenditure | -0.30 | (-0.31 to -0.29) | -1.65 | (-1.73 to -1.58) | -3.31 | (-3.56 to -3.06) |
| ***Revenue*** |  |  |  |  |  |  |
| Income tax revenue | 0.29 | (0.25 to 0.35) | 1.78 | (1.49 to 2.11) | 3.98 | (3.32 to 4.73) |
| GST revenue (including tobacco sales tax) | 0.50 | (0.43 to 0.58) | 1.63 | (1.4 to 1.88) | 3.01 | (2.57 to 3.49) |
| Tobacco excise revenue | -5.24 | (-6.16 to -4.24) | -10.35 | (-12.26 to -8.24) | -13.56 | (-16.39 to -10.53) |
| **Net Government position**<br>($\sum \text{revenue} - \sum \text{expenditure}$) | -3.96 | (-4.8 to -3.07) | -4.54 | (-6.35 to -2.65) | -1.89 | (-4.74 to 1.01) |
| **Citizen perspective** |  |  |  |  |  |  |
| Population income after tax | 0.82 | (0.68 to 0.97) | 4.96 | (4.15 to 5.88) | 11.07 | (9.24 to 13.15) |
| Savings from cessation (reduced tobacco expenditure) | 11.35 | (13.35 to 9.19) | 22.41 | (26.55 to 17.83) | 29.36 | (35.49 to 22.8) |
| Population income after tax + savings from cessation | 12.17 | (9.95 to 14.2) | 27.38 | (22.52 to 31.78) | 40.45 | (33.18 to 47.3) |

Note: 2020 NZ\$ converted to 2021 US\$ using NZ-US OECD purchasing power parity of 1.4684.
† i.e., includes estimate to left, as cumulative over time.

## DISCUSSION

Our modelling suggests that the Smokefree Aotearoa 2025 Action Plan recently passed into law by the A/NZ Government is likely to produce substantial economic benefits in addition to the previously calculated^9^ health and health equity benefits. Under this scenario, the population would benefit from a cumulative gain in post-tax income of US$1.8 billion by 2050. Factoring in consumer savings on tobacco expenditure, leads to a cumulative increase of US$31 billion in total disposable income by 2050. Our estimates are consistent with a large body of evidence documenting the detrimental impact of tobacco spending on household budgets, particularly for the most disadvantaged socio-economic categories.^17^ An analysis of A/NZ census data has estimated that among low-income households with at least one member who smokes, up to 14% of the non-housing budget was spent on tobacco.^18^ Similar findings have been reported in other high-income countries^19^ as well as low-^20,21^ and middle-income countries.^22,23^ A recent modelling study evaluating the economic loss attributable to cigarette smoking in the US estimated the total loss in annual population income in 2020 at US$735.1 billion.^3^ In A/NZ Moreover, smoking is strongly concentrated among Indigenous Māori and people on low incomes ^24^, therefore our estimated increases in disposable income would represent a pro-equity income transfer.

From a government perspective, the picture is more mixed. A/NZ has one of the world’s highest tobacco excise tax. In 2021, the pack price of 20 Marlboro cigarettes was NZ$36.9 (US$25) with excise tax and GST representing 70% of the price. Consequently, the Government foregoes considerable excise tax revenue under the endgame policy scenario. Despite clear direct financial benefits (from reduced health expenditure and increases to both GST from higher population income and income tax revenue), the Government’s long-term net position remains negative in our primary analysis with a fixed age eligibility to superannuation benefit, due to the decline of excise tax revenue (Figure 1). This decrease in revenue is a logical consequence of successfully reducing smoking prevalence and was identified in the Regulatory Impact Statement preceding the legislation.^8^

Tobacco excise tax revenue also decreased under BAU – this is again a logical result of the underlying decreasing trend in smoking prevalence.^24^ The endgame policy simply accelerates the rate of decline of this revenue source. In 2019/20, tobacco tax revenue was about 1.7% of annual A/NZ Government revenue,^13^ which is relatively small compared to annual variation in government revenue arising from typical macroeconomic fluctuations and natural hazards that have impacted A/NZ in recent decades (major earthquakes, major storms and the Covid-19 pandemic).

Previous analyses that examined the impact of reducing smoking in the US to 10.4% (the estimated impact of the Institute of Medicine (IOM) recommended policy package) or to 5.7% (a hypothetical high impact scenario) by 2025 on a range of economic outcomes also found the high impact scenario would reduce state government tobacco tax revenue on average by 2.5% due to the greater decline in cigarette sales, while the IOM policy package would produce a 0.5% increase by raising the tax rate.^25^

Through its world first tobacco endgame legislation, the A/NZ Government has prioritised health and equity over government revenue with three of the five political parties in the Parliament also fully supporting it (and the main opposition party still supportive of the denicotinisation policy). However, in countries where economic priorities are perceived as more important,^5^ the cost to government revenue may present a major potential barrier to progressing a tobacco endgame. This may be particularly challenging in low- and middle-income countries, which may not yet be experiencing the full adverse health and economic impacts of the tobacco epidemic while collecting tobacco tax revenue from growing tobacco sales. Our modelling, although specific to the A/NZ context, assumes that a tobacco endgame strategy is likely to result in large economic benefits for the population and that revenue foregone by governments is not lost but rather re-transferred to the population. A tobacco endgame may also address a key ethical challenge that tobacco taxation can pose in terms of contributing to financial hardship among low-income households where smoking persists.

Our study estimated an increase in government spending on aged pensions over time due to the reduction in premature mortality from tobacco-related disease – assuming a fixed age of eligibility to universal Government superannuation. Tobacco companies have previously attempted to sell the ‘financial benefits’ of smoking to governments in the form of reduced expenditure on aged pensions due to the reduced life expectancy.^26^ However, since increasing life expectancy and health is a societal (and government) goal, increased financial costs associated with such health benefits in the form of government superannuation/aged pensions should not be a determining factor in government-decision making regarding policies that have life extending benefits.^27^ Nevertheless, estimating these impacts can assist the A/NZ government to plan appropriately as the country becomes smokefree.

Acknowledging current Stats NZ projections of a larger and older working-age population in A/NZ, our sensitivity analysis scenario using a dynamic “retirement age” suggests that the government can achieve a positive net fiscal position despite the losses in excise tax revenue associated with the endgame policy package. This ‘recovery’ occurs only 14-years after the introduction of the policy and with minor incremental increases to the age of superannuation entitlement – from 65 years in 2020 to 65.78 years by 2050. Such a policy is consistent with changes being implemented in other similar countries. For example, Australia is currently gradually increasing the eligibility age for the aged pension from 65 years at 30 June 2017 to 67 years on 1 July 2023.^28^ Nevertheless, our dynamic scenario is just that – a scenario. No Government would change the retirement age and age of eligibility for universal superannuation benefits by such small increments per annum. Our purpose was to demonstrate how increased healthiness of the population might manifest as one way for society to adjust.

This study used a tobacco policy model ranked top of 15 such models internationally.^29^ However, it has several limitations. Our modelling attempts to estimate what the future might look like and so has many uncertainties. To help capture these uncertainties we applied a probabilistic sensitivity analysis approach to the key parameters in our simulation (in line with the recommendations for best practice^30^) to generate uncertainty ranges for the outputs. The model also relies on multiple assumptions and is therefore exposed to the limitations associated with those assumptions (see full discussion of model limitations in the main manuscript and supplementary material of Ait Ouakrim et al^9^). In our model, ‘retail reduction’ was specified as a 95% reduction in the number of tobacco retailers, based on earlier pre-legislation plans. However, the final version of the ‘smokefree bill’ adopted a minimum 90% reduction in tobacco retail stores which translates to a maximum of 600 retail outlets. This might slightly over-estimate the projected economic benefits and, conversely, under-estimate the expenditure associated with the endgame policy package.

Our estimate of the net government position should be interpreted with caution as it is only limited to the macroeconomic outputs considered in the model. For example, the model did not take into account the health benefits (and subsequent economic dividends) that the endgame might produce as a result of lower population exposure to second hand smoke.^31,32^ Similarly, we did not take into account the many Government tax revenue sources (such as corporate tax, taxes on payroll and workforce, tax on production etc.) that are likely to increase and benefit from a healthier and more productive population. Finally, our model did not account for the out-of-pocket health expenditure savings for citizens that would result from lower disease and treatment burden associated with quitting and lower uptake of smoking.

Smoking imposes intangible detrimental effects on people and society (for example the psychological pain associated with chronic addiction, tobacco-related disease and the prospect of premature death). These effects impact quality of life and productivity in the formal and informal economies. But they are hard to value quantitatively and our model does not take them into account. Consequently, our estimates of both the economic benefits of the tobacco endgame are likely to be under-estimates.

## CONCLUSION

Our study estimated the expected economic impacts of the Smokefree Aotearoa 2025 Action Plan, demonstrating economic benefits for the A/NZ population, and modest impacts on government revenue and expenditure related to the reduction in tobacco tax and increases in aged pensions due to increased life expectancy. Such costs are relatively small compared to other macroeconomic fluctuations, and can be anticipated and planned for. These costs could be offset by future increases in labour force and the proportion of 65+ year olds working in the formal economy.

## Data Availability

All data produced in the present study are available upon reasonable request to the authors

## References

1. Ezzati M, Lopez AD, Rodgers A, Vander Hoorn S, Murray CJ. Selected major risk factors and global and regional burden of disease. the lancet 2002; 360(9343): 1347–60.

2. Goodchild M, Nargis N, d’Espaignet ET. Global economic cost of smoking-attributable diseases. Tobacco control 2018; 27(1): 58–64.

3. Nargis N, Hussain AG, Asare S, et al. Economic loss attributable to cigarette smoking in the USA: an economic modelling study. The Lancet Public Health 2022; 7(10): e834–e43.

4. Malone RE. The race to a tobacco endgame. BMJ Publishing Group Ltd; 2016. p. 607–8.

5. Moon G, Barnett R, Pearce J, Thompson L, Twigg L. The tobacco endgame: the neglected role of place and environment. Health & place 2018; 53: 271–8.

6. Puljević C, Morphett K, Hefler M, et al. Closing the gaps in tobacco endgame evidence: a scoping review. Tobacco Control 2022; 31(2): 365–75.

7. Parliament NZ. Smokefree Environments and Regulated Products (Smoked Tobacco) Amendment Act 2022 (2022/79). 2022. https://www.parliament.nz/en/pb/bills-and-laws/bills-proposed-laws/document/BILL_125245/smokefree-environments-and-regulated-products-smoked-tobacco.

8. Ministry of Health. Regulatory Impact Statement: Smokefree Aotearoa 2025 Action Plan. Wellington: Ministry of Health 2021. 2021. https://www.health.govt.nz/about-ministry/information-releases/regulatory-impact-statements/regulatory-impact-statement-smokefree-aotearoa-2025-action-plan (accessed 5/02/2023).

9. Ait Ouakrim D, Wilson T, Waa A, et al. Tobacco endgame intervention impacts on health gains and Māori:non-Māori health inequity: a simulation study of the Aotearoa/New Zealand Tobacco Action Plan. Tobacco Control 2023: tc-2022-057655.

10. Blakely T, Cobiac LJ, Cleghorn CL, et al. Health, health inequality, and cost impacts of annual increases in tobacco tax: multistate life table modeling in New Zealand. PLoS medicine 2015; 12(7): e1001856.

11. Pearson AL, Cleghorn CL, van der Deen FS, et al. Tobacco retail outlet restrictions: health and cost impacts from multistate life-table modelling in a national population. Tob Control 2016.

12. Petrović-van der Deen FS, Blakely T, Kvizhinadze G, Cleghorn CL, Cobiac LJ, Wilson N. Restricting tobacco sales to only pharmacies combined with cessation advice: a modelling study of the future smoking prevalence, health and cost impacts. Tobacco control 2019; 28(6): 643–50.

13. Treasury NZ. Financial statement of the Government of New Zealand (year ending June 2021). Available from: 2021.

14. Murray CJ, Ezzati M, Lopez AD, Rodgers A, Vander Hoorn S. Comparative quantification of health risks: conceptual framework and methodological issues. Population health metrics 2003; 1(1): 1–20.

15. Stats New Zealand. National labour force projections: 2020(base)–2073. 2021. https://www.stats.govt.nz/information-releases/national-labour-force-projections-2020base-2073 (accessed 9/02/2023.

16. Blakely T, Moss R, Collins J, et al. Proportional multistate lifetable modelling of preventive interventions: concepts, code and worked examples. International Journal of Epidemiology 2020; 49(5): 1624–36.

17. National Cancer Institute, World Health Organization. The Economics of Tobacco and Tobacco Control. National Cancer Institute Tobacco Control Monograph 21. NIH Publication No. 16-CA-8029A. Bethesda, MD: U.S. Department of Health and Human Services, National Institutes of Health, National Cancer Institute; and Geneva, CH: World Health Organization. 2016. https://cancercontrol.cancer.gov/brp/tcrb/monographs/21/docs/m21_complete.pdf.

18. Thomson GW, Wilson N, O’dea D, Reid P, Howden-Chapman P. Tobacco spending and children in low income households. Tobacco Control 2002; 11(4): 372–5.

19. Nyakutsikwa B, Britton J, Langley T. The effect of tobacco and alcohol consumption on poverty in the United Kingdom. Addiction 2021; 116(1): 150–8.

20. Husain MJ, Datta BK, Virk-Baker MK, Parascandola M, Khondker BH. The crowding-out effect of tobacco expenditure on household spending patterns in Bangladesh. PloS one 2018; 13(10): e0205120.

21. Nguyen N-M, Nguyen A. Crowding-out effect of tobacco expenditure in Vietnam. Tobacco Control 2020; 29(Suppl 5): s326–s30.

22. San S, Chaloupka FJ. The impact of tobacco expenditures on spending within Turkish households. Tobacco control 2016; 25(5): 558–63.

23. Wang H, Sindelar JL, Busch SH. The impact of tobacco expenditure on household consumption patterns in rural China. Social science & medicine 2006; 62(6): 1414–26.

24. Ministry of Health. Annual Data Explorer 2021/22: New Zealand Health Survey Tobacco Use. 2022. <https://minhealthnz.shinyapps.io/nz-health-survey-2021-22-annual-data-explorer/>.

25. Bearman PS, Neckerman KM, Wright L. After tobacco: what would happen if Americans stopped smoking?: Columbia University Press; 2011.

26. Ross H. Critique of the Philip Morris study of the cost of smoking in the Czech Republic. Nicotine & tobacco research 2004; 6(1): 181–9.

27. Barendregt JJ, Bonneux L, van der Maas PJ. The health care costs of smoking. New England Journal of Medicine 1997; 337(15): 1052–7.

28. Services AGDoS. Seniors Benefits & Payments - Age Pension. 2023. https://www.dss.gov.au/seniors/benefits-payments/age-pension (accessed 15 February 2023.

29. Huang V, Head A, Hyseni L, et al. Identifying best modelling practices for tobacco control policy simulations: a systematic review and a novel quality assessment framework. Tobacco control 2022.

30. Briggs AH, Weinstein MC, Fenwick EA, Karnon J, Sculpher MJ, Paltiel AD. Model parameter estimation and uncertainty analysis: a report of the ISPOR-SMDM Modeling Good Research Practices Task Force Working Group–6. Medical decision making 2012; 32(5): 722–32.

31. Callinan JE, Clarke A, Doherty K, Kelleher C. Legislative smoking bans for reducing secondhand smoke exposure, smoking prevalence and tobacco consumption. Cochrane Database Syst Rev 2010; (4).

32. Christensen TM, Møller L, Jørgensen T, Pisinger C. The impact of the Danish smoking ban on hospital admissions for acute myocardial infarction. European journal of preventive cardiology 2014; 21(1): 65–73.

33. Stats New Zealand. Earnings for people in paid employment by region, sex, age groups and ethnic groups. https://nzdotstat.stats.govt.nz/wbos/index.aspx?_ga=2.59390884.807928993.1671346727-697363774.1671144874#

34. Blakely T, Sigglekow F, Irfan M, et al. Disease-related income and economic productivity loss in New Zealand: A longitudinal analysis of linked individual-level data. PLoS medicine 2021; 18(11): e1003848.

35. Drope J, Hamill S, Chaloupka F, et al. The Tobacco Atlas. 2022. https://tobaccoatlas.org/.

