## Supplementary Material for "Economic effects of a country-level tobacco endgame strategy: a modelling study"

**Figure S1. Tobacco and vaping life history Markov model**

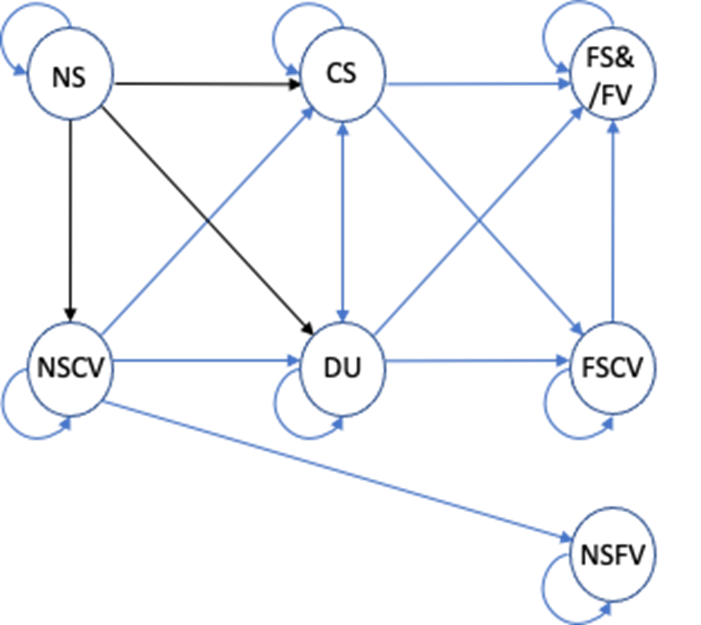

NS = never smoker, CS = current smoker, FS & FV = former smoker and former vaper; NSCV = never smoker current vaper; DU = dual user; FSCV = former smoker current vaper; NSFV = never smoker former vaper.

**Table S1. Intervention input parameters***

| **Parameter** | **Description** |
| --- | --- |
| ***Denicotinisation*** | |
| NS🡪 CS (age 20 only)  NS🡪 DU (age 20 only) | 90% (SD 5%) of BAU initiation at age 20 by five years after implementation (X= Beta (32.4, 3.6), median 90.7%, 95% UI: 78.5% to 97.4%.). Implemented as 1 – (1-X)^(t/5) scalar applied to the BAU initiation rates in years t (1 to 5) after introduction of the policy, then held at 1 – X% thereafter. |
| CS🡪 FSFV  CS🡪 FSCV  DU🡪 FSFV  DU🡪 FSCV | Using an expert knowledge elicitation (see Appendix D), the reduction in smoking prevalence five years after the low nicotine policy compared to BAU in five years, due to quitting or switching to vaping, was mean 84.4% (SD 7.84%, X=Beta (17.78, 3.19), median 85.9%, 95%UI: 67.1% to 96.3%). Implementation was as 1 – (1 – X)^(t/5) scalar applied to BAU CS and DU prevalence, where t is the 1 to 5 years after intervention. For the sixth and subsequent years, the transition probabilities were twice those in BAU (due to an ongoing higher NCR, given non-addictive levels of nicotine in tobacco). |
| NS🡪NSCV | No change. |
| ***Denicotinisation plus Mass media*** | |
| NS🡪 CS (age 20 only) | As above for low nicotine. |
| NS🡪 DU (age 20 only) | As above for low nicotine. |
| NS🡪 NSCV (age 20 only) | No change. |
| CS🡪 FSFV  CS🡪 FSCV  DU🡪 FSFV  DU🡪 FSCV | As above for low nicotine from year 1 to 5 +  twice the absolute contribution of the routine media/Quitline campaign added to background net cessation (i.e. 1.055% × 2 = 2.1%)^1^  Subsequent years: transition to quitting or vaping were twice those in BAU |
| ***Retail outlet restriction to about 300 outlets (about 5% of current outlets; assumed supply of e-cigarettes reduces commensurately) ^†^*** | |
| NS🡪 CS | As per the increase in cessation probabilities (CS🡪FSFV, etc, below), we reduced the initiation rate by X= Beta (23.4, 97.2), median 19.2%, 95%UI: 12.9% to 26.9%. Applies in 2023 onwards (as youth contemplating initiating in the future confront lesser retail availability as well). |
| NS🡪 DU | As above for NS🡪CS. |
| CS🡪 FSFV  CS🡪 FSCV  DU🡪 FSFV  DU🡪 FSCV | As a low estimate of one-off quitting, we used that from studies modelling reducing retail outlets in terms of increased travel costs ^2^: a reduction in the prevalence of 15.6% for Māori, and 16.0% for non-Māori – or 15.8% overall.  As a high estimate, we used that from the NZ ITC study where – in response to a question whether they would quit in response to a 95% reduction in retail outlets - 23.0% said they would quit (half quitting 🡪 FSFV, half switching to FSCV).^3^  Placing the mean at 19.4% (average of above 15.8% and 23%) and using 15.8% and 23% as one SD either side of the mean (SD = 3.6%), we parameterised the one-off increase in smoking net cessation as X=Beta (23.4, 97.2), median 19.2% (i.e., percentage point increase), 95%UI: 12.9% to 26.9%. Note this increase was on top of BAU transition probabilities and halved over CS🡪FSFV and CS🡪FSCV and halved over DU🡪FSFV and DU🡪FSCV. E.g., if the CS🡪FSFV was 5%, the intervention CS🡪FSFV transition probability was 5% + (1-5%) × 0.5 × X%.  This effect was in the year of intervention only– in years after the retail outlet restriction, the transition probabilities out of CS and DU reverted to BAU. |
| NS🡪NSCV | Unchanged |
| ***Tobacco-free generation*** | |
| Smoking initiation rate (NS🡪 CS; occurs only at age 20) | For two reasons, a tobacco-free generation proposal will not immediately achieve zero uptake at age 20; 1) our model for parsimony assumes all uptake at age 20, but the minimum legal age of purchasing is 18 years; 2) social supply will allow some young people to keep initiating. We therefore assumed that initiation at age 20 in our model (essentially an average of all initiation by [say] age 25) will asymptote to a mean of X=10% (SD 5%) of BAU in 10 years (Beta (3.6, 32.4), median 9.3%, 95%UI: 2.6% to 21.5%), with the scalar of BAU initiation rate of X^(t/10) for t = 1 to 10 years after the tobacco-free generation policy is implemented, then X of BAU initiation thereafter. |
| NS🡪 DU | As above for NS🡪CS. |
| NS🡪 NSCV | Unchanged† |
| ***Combined: Denicotinisation + media +retail + tobacco-free generation*** | |
| NS🡪 CS (age 20 only) | Cumulative impact. If the % reduction in initiation in year t for denicotinisation, retail and tobacco-free was A%, B%, and C%, then the reduction in the combined intervention was 1 – (1-A)(1-B)(1-C). |
| NS🡪 DU (age 20 only) | As above for NS🡪CS. |
| CS🡪 FSFV  CS🡪 FSCV  DU🡪 FSFV  DU🡪 FSCV | Cumulative impact. If the % increase in quitting or switching in year t for denicotinisation, media and retail was A% and B%, then the increase in the combined intervention was 1 – (1-A)(1-B)(1-C). |
| NS🡪 NSCV | Unchanged. |

*reproduced from Ait Ouakrim et al.^4^

**Table 2: Projected changes in cumulative expenditure and revenue due to the Aotearoa-New Zealand’s tobacco endgame strategy compared to BAU (2021 PPP US$ billions; undiscounted)**

| Revenue/expenditure items | by 2030 | | by 2040† | | by 2050† | |
| --- | --- | --- | --- | --- | --- | --- |
| **Government perspective** | **Estimate** | **95% UI** | **Estimate** | **95%** | **Estimate** | **95%** |
| ***Expenditure*** |  |  |  |  |  |  |
| Health system | 0.23 | (0.28 to 0.18) | 1.12 | (1.38 to 0.88) | 2.45 | (3.13 to 1.85) |
| Superannuation expenditure | 0.04 | (0.03 to 0.05) | 0.58 | (0.45 to 0.7) | 2.42 | (1.9 to 2.94) |
| ***Revenue*** |  |  |  |  |  |  |
| Income tax revenue | 0.06 | (0.05 to 0.08) | 0.47 | (0.37 to 0.56) | 1.24 | (1 to 1.48) |
| GST revenue (including tobacco sales tax) | 0.46 | (0.37 to 0.54) | 1.21 | (0.98 to 1.42) | 2.08 | (1.66 to 2.48) |
| Tobacco excise revenue | -6.44 | (-7.55 to -5.22) | -14.56 | (-17.27 to -11.55) | -21.49 | (-26.19 to -16.5) |
| ***Net Government position***  ***(revenue –expenditure)*** | -5.72 | (-6.72 to -4.63) | -12.34 | (-14.73 to -9.68) | -18.13 | (-22.34 to -13.58) |
| **Citizen perspective** |  |  |  |  |  |  |
| Population income after tax | 0.18 | (0.14 to 0.21) | 1.30 | (1.04 to 1.55) | 3.44 | (2.79 to 4.11) |
| Savings from cessation (reduced tobacco expenditure) | 13.94 | (16.35 to 11.29) | 31.54 | (37.39 to 25.02) | 46.52 | (56.71 to 35.72) |
| Population income after tax + savings from cessation | 14.11 | (11.44 to 16.56) | 32.83 | (26.13 to 38.82) | 49.96 | (38.68 to 60.49) |

Note: 2020 NZ$ converted to 2021 US$ using NZ-US OECD purchasing power parity of 1.4684.

† i.e., includes estimate to left, as cumulative over time.

**Table 3 Projected changes in cumulative expenditure and revenue due to the Aotearoa-New Zealand tobacco endgame strategy compared to BAU using dynamic retirement age (2021 PPP USD billions; undiscounted)**

| Revenue/expenditure items | by 2030 | | by 2040† | | by2050† | |
| --- | --- | --- | --- | --- | --- | --- |
| **Government perspective** | **Estimate** | **95% UI** | **Estimate** | **95%** | **Estimate** | **95%** |
| ***Expenditure*** |  |  |  |  |  |  |
| Health system | 0.23 | (0.28 to 0.18) | 1.12 | (1.38 to 0.88) | 2.45 | (3.13 to 1.85) |
| Superannuation expenditure | -0.39 | (-0.39 to -0.38) | -2.59 | (-2.71 to -2.47) | -6.24 | (-6.75 to -5.72) |
| ***Revenue*** |  |  |  |  |  |  |
| Income tax revenue | 0.38 | (0.32 to 0.45) | 2.82 | (2.36 to 3.34) | 7.65 | (6.39 to 9.09) |
| GST revenue (including tobacco sales tax) | 0.63 | (0.53 to 0.72) | 2.46 | (2.11 to 2.83) | 5.48 | (4.69 to 6.38) |
| Tobacco excise revenue | -6.44 | (-7.55 to -5.22) | -14.56 | (-17.27 to -11.55) | -21.49 | (-26.19 to -16.5) |
| ***Net Government position***  ***(revenue – expenditure)*** | -4.82 | (-5.83 to -3.73) | -5.55 | (-8.14 to -2.85) | 0.38 | (-4.48 to 5.28) |
| **Citizen perspective** |  |  |  |  |  |  |
| Population income after tax | 1.05 | (0.88 to 1.24) | 7.83 | (6.55 to 9.29) | 21.28 | (17.76 to 25.29) |
| Savings from cessation (reduced tobacco expenditure) | 13.94 | (16.35 to 11.29) | 31.54 | (37.39 to 25.02) | 46.52 | (56.71 to 35.72) |
| Population income after tax + savings from cessation | 14.99 | (12.27 to 17.44) | 39.38 | (32.43 to 45.67) | 67.85 | (55.63 to 79.49) |

Note: 2020 NZ$ converted to 2021 US$ using NZ-US OECD purchasing power parity of 1.4684.

† i.e., includes estimate to left, as cumulative over time.

**Figure S1: Estimated annual differences in revenue and expenditure (2021 US$; undiscounted) between the tobacco endgame scenario and BAU**

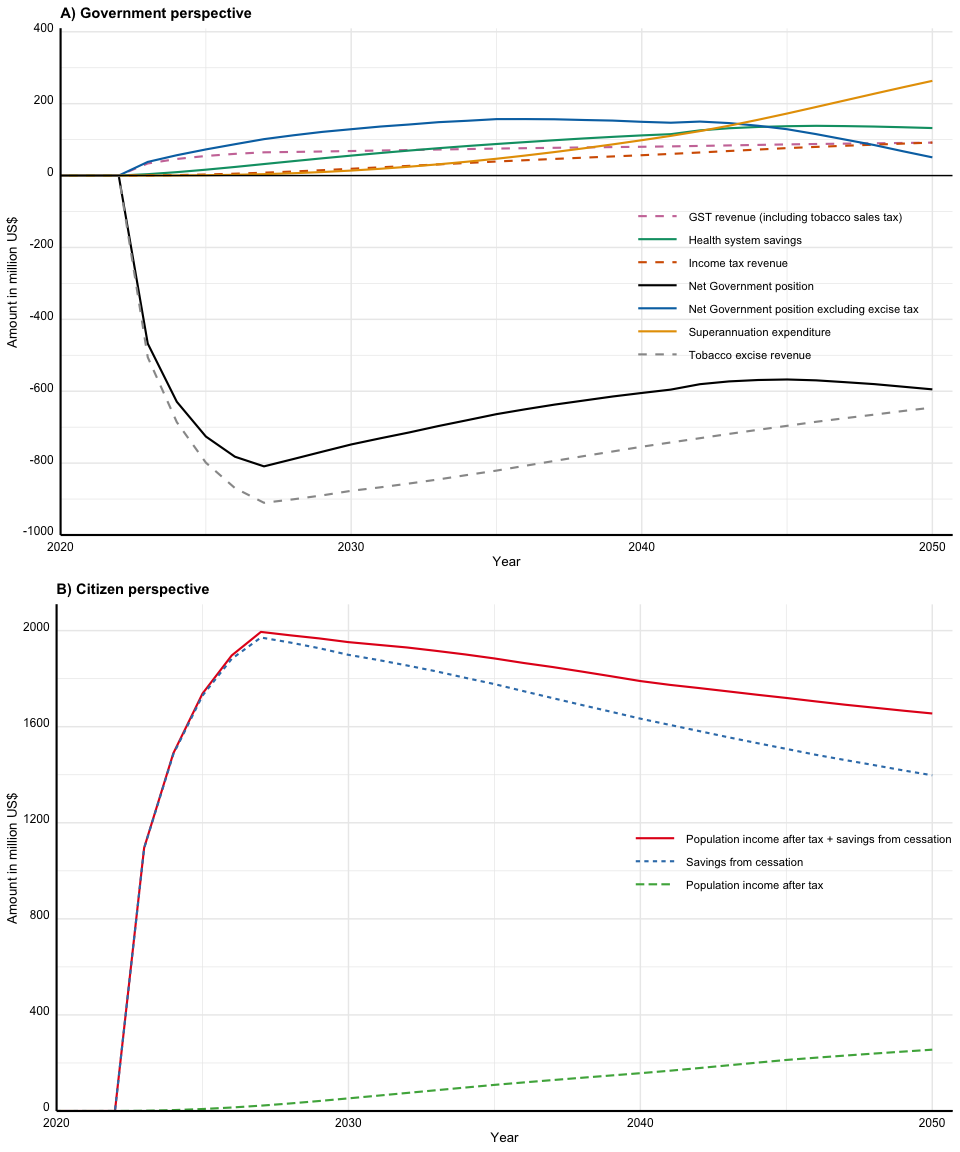

**Figure S2: Estimated annual differences in revenue and expenditure (2021 PPP US$; undiscounted) between tobacco endgame scenario with dynamic retirement age and BAU (Government perspective)**

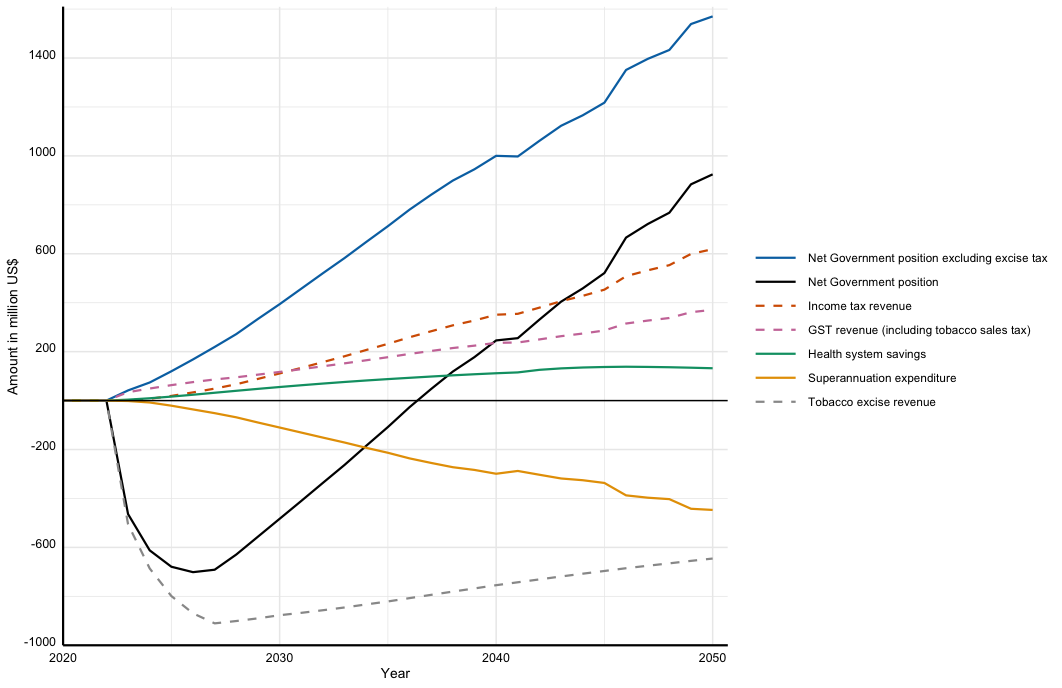

**References**

1.	Nghiem N, Cleghorn CL, Leung W, et al. A national quitline service and its promotion in the mass media: modelling the health gain, health equity and cost–utility. *Tobacco Control* 2017; **27**(4): 434-41.

2.	Petrovic-Van Der Deen FS, Blakely T, Kvizhinadze G, Cleghorn CL, Cobiac LJ, Wilson N. Restricting tobacco sales to only pharmacies combined with cessation advice: A modelling study of the future smoking prevalence, health and cost impacts. *Tobacco Control* 2019; **28**(6): 643-50.

3.	Edwards R, Johnson E, Hoek J, et al. The Smokefree 2025 Action Plan: key findings from the ITC New Zealand (EASE) project. In: Expert PH, editor. Public Health Expert. Wellington: University of Otago; 2021.

4.	Ait Ouakrim D, Wilson T, Waa A, et al. Tobacco endgame intervention impacts on health gains and Māori:non-Māori health inequity: a simulation study of the Aotearoa/New Zealand Tobacco Action Plan. *Tobacco Control* 2023: tc-2022-057655.
